# Transcriptome and Functions of Granulocytic Myeloid-Derived Suppressor Cells Determine their Association with Disease Severity of COVID-19

**DOI:** 10.1101/2021.03.26.21254441

**Authors:** Matthew J. Dean, Juan B. Ochoa, Maria Sanchez-Pino, Jovanny Zabaleta, Jone Garai, Luis Del Valle, Dorota Wyczechowska, Lyndsey Buckner, Phaethon Philbrook, Rinku Majumder, Richard Vander Heide, Logan Dunkenberger, Ramesh Thylur, Robert Nossaman, W. Mark Roberts, Andrew Chapple, Jack Collins, Brian Luke, Randall Johnson, Hari Koul, Christopher A. Rees, Claudia R. Morris, Julia Garcia-Diaz, Augusto C. Ochoa

## Abstract

COVID-19 ranges from asymptomatic in 35% of cases to severe in 20% of patients. Differences in the type and degree of inflammation appear to determine the severity of the disease. Recent reports show an increase in circulating monocytic-myeloid-derived suppressor cells (M-MDSC) in severe COVID 19, that deplete arginine but are not associated with respiratory complications. Our data shows that differences in the type, function and transcriptome of Granulocytic-MDSC (G-MDSC) may in part explain the severity COVID-19, in particular the association with pulmonary complications. Large infiltrates by Arginase 1^+^ G-MDSC (Arg^+^G-MDSC), expressing NOX-1 and NOX-2 (important for production of reactive oxygen species) were found in the lungs of patients who died from COVID-19 complications. Increased circulating Arg^+^G-MDSC depleted arginine, which impaired T cell receptor and endothelial cell function. Transcriptomic signatures of G-MDSC from patients with different stages of COVID-19, revealed that asymptomatic patients had increased expression of pathways and genes associated with type I interferon (IFN), while patients with severe COVID-19 had increased expression of genes associated with arginase production, and granulocyte degranulation and function. These results suggest that asymptomatic patients develop a protective type I IFN response, while patients with severe COVID-19 have an increased inflammatory response that depletes arginine, impairs T cell and endothelial cell function, and causes extensive pulmonary damage. Therefore, inhibition of arginase-1 and/or replenishment of arginine may be important in preventing/treating severe COVID-19.

## INTRODUCTION

COVID-19 disease ranges from asymptomatic (35% of cases) to severe requiring treatment in intensive care units (*https://www.cdc.gov/coronavirus/2019-ncov/hcp/clinical-guidance-management-patients.html*). Approximately 20% of hospitalized COVID-19 patients develop hypoxemia, respiratory difficulty, hypercoagulation and end organ damage that may result in death. These clinical manifestations are similar to other coronavirus infections including SARS-CoV-1 and MERS-CoV. The pathophysiology is primarily associated with an over-active inflammatory response manifested by a cytokine release syndrome, a surge in granulocytes and decreased lymphocytes^1,2^. The mechanism(s) that regulate these events are unclear. Genomic studies of peripheral blood suggested in increase genomic signatures associated with neutrophil functions in COVID-19 patients^3^. Early reports also showed that increased neutrophils/lymphocyte ratios were associated with poor outcomes^4,5^. More recently, Agrati *et al*^*6*^, Reizine *et al* ^7^ and Falk-Jones *et al* ^8^ showed increased circulating monocytic -myeloid derived suppressor cells (M-MDCS) able to deplete arginine and inhibit T cell proliferation. MDSC have been best studied in cancer, but also described in chronic infections, autoimmunity, asthma and trauma^9-11^. MDSC express high arginase 1 (Arg1) that metabolizes arginine to ornithine and urea, effectively depleting this amino acid from the microenvironment. Arginine depletion inhibits T cell receptor signaling resulting in T cell dysfunction^12,13^. It also impairs nitric oxide production and increases endothelial cell dysfunction promoting intravascular coagulation ^14,15^. Arginine depletion also increases the production of reactive oxygen species (ROS) which can damage infiltrated organs and exacerbate inflammation ^16^. We compared the type, function and transcriptomic signature of MDSC in COVID-19 patients to better understand their effect on arginine levels, T cell function and respiratory complications. Our results showed a major increase in circulating *granulocytic*-MDSC expressing high levels of arginase-1 (Arg^+^G-MDCS). Large accumulations of Arg^+^G-MDSC expressing NOX1 and NOX2 infiltrated the lungs of patients who died from severe COVID-19 complications. High numbers of Arg^+^G-MDSC in circulation depleted arginine in plasma, decreased T cell receptor zeta chain (CD3ς) and increased markers of endothelial cell dysfunction. RNAseq studies demonstrated contrasting transcriptomes, where G-MDSC from asymptomatic COVID-19 patients had increased expression of type I IFN genes and pathways, while G-MDSC from severe COVID-19 patients had instead increased expression of genes associated with granulocyte functions and degranulation. These data support the concept that Arg^+^G-MDSC may play a significant role in the pathophysiology of COVID-19.

## RESULTS

### Study participants

The study was conducted under LSU IRB protocol #20-053 and Ochsner Medical Center IRB protocol 21015-101C. All participants were consented prior to inclusion in the study. Peripheral blood from 24 severe, 26 convalescent and 5 asymptomatic COVID-19 patients and 15 healthy (COVID-19 negative) controls were used for these studies (Supplemental Table 1). Severity of the disease was determined following WHO classification. All severe COVID-19 patients were being treated in the ICU at the time of sample collection. Treatments included remdesivir, systemic corticosteroids, antibiotics and anticoagulants; with convalescent sera or monoclonal antibodies used in 3 cases. Only 50% of severe COVID-19 patients were on ventilator support. Samples were collected within the first three weeks of admission to ICU (mean= 17.6 days; range 2-77 days). Asymptomatic and convalescent patients were ambulatory. Average age varied from 43.4 in asymptomatic patients to 67.8 in severe COVID-19 patients. Average Body Mass Index (BMI), a risk factor for severity, ranged from 25 in healthy controls to 32.5 in severe COVID-19 patients. Tissues from 10 patients who died from COVID-19 complications were tested for inflammatory infiltration.

### Dysregulated Immune Profiles in Severe COVID-19

Flow cytometry of peripheral blood (Figure 1) showed a significant decrease in CD3+ T cells (1A) (both CD4 and CD8 – data not shown) and CD16+ NK cells (1C), no significant changes in CD14+ monocytes (1D), and a moderate but significant increase in CD66b+ granulocytes (1B). However, CD66b granulocyte: CD3 T cells (1E) and CD16 NK (1F) cell ratios were dramatically increased in severe COVID-19 patients. Separation of peripheral blood mononuclear cells (PBMC) over ficoll-hypaque, which eliminates high density granulocytes, further revealed a significant increase in G-MDSC in severe COVID-19 patients ranging from 2.3% −76% (mean=29.3%) (1G), which was >10 fold higher than normal controls (mean=2.5%), asymptomatic and convalescent patients (mean=1.2%). In 62% (14/24) of severe COVID-19 patients G-MDSC were >20% of PBMC following ficoll-histopaque separation, i.e. > 4 standard deviations above the mean of normal controls and convalescent patients. G-MDSC levels did not differ between severe COVID-19 patients needing ventilator support, the presence of pneumonia, nor differences in medications (data not shown). Monocytic-MDSC (1H) were also moderately increased, but not as significantly as G-MDSC. Comparison of the flow cytometry results using Log Density Plots (1I) showed significantly higher CD66b/CD3 (p=6.8 × 10^−8^), G-MDSC/CD3 (4.07 × 10^−12^) ratios and percent G-MDSC (1.68 × 10^−16^) in severe COVID-19 patients compared to other patients. These major differences suggest that these ratios and the percent G-MDSC may help identify patients progressing toward a severe form of COVID-19.

**Figure 1.**
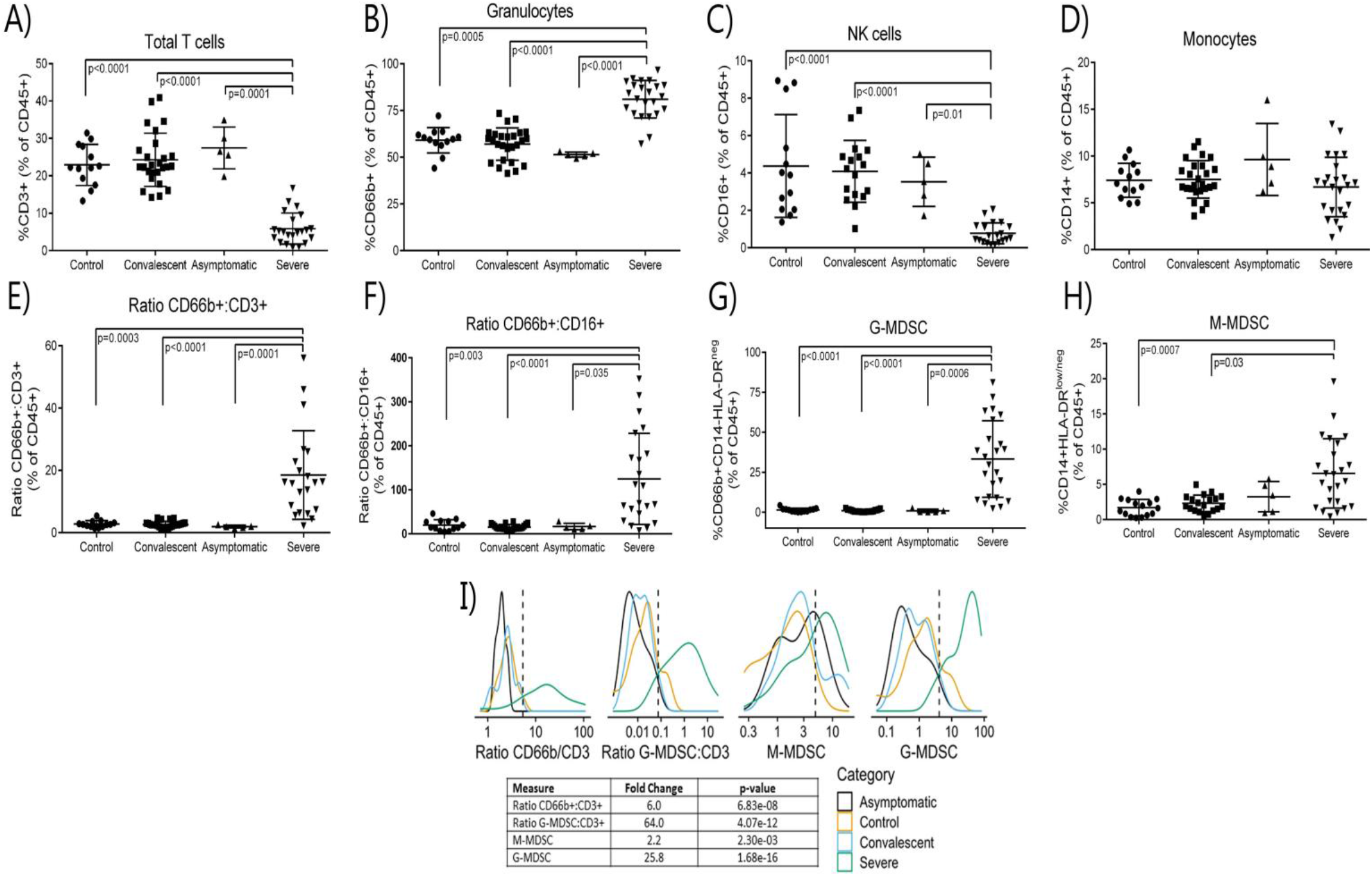
Leukocyte Subpopulations including MDSC in Peripheral Blood of COVID-19 Patients. Whole blood and PBMCs from all participants were analyzed by flow cytometry. Percent **A)** T cells, **B)** granulocytes, **C)** NK cells, and **D)** monocytes in whole blood. **E)** CD66b/CD3 and **F)** CD66b/CD16 ratios in whole blood. Percent **G)** G-MDSC and **H)** M-MDSC in PBMC isolated over ficoll-hypaque; **I)** Gaussian Kernel Density using a log10-scaled density of observed values for CD66b:CD3 ratios, G-MDSC:CD3 ratios, M-MDSC, G-MDSC. The vertical, dashed line represents the thresholds obtained in the sensitivity/specificity analysis providing the maximum separation between severe COVID-19 patients and other groups of patients or healthy controls.

### G-MDSC infiltration of the lungs

A frequent complication of severe COVID-19 is hypoxemia and acute respiratory distress. We tested whether G-MDSC might play a role in the pathophysiology of this complication. Lung autopsy samples from ten patients who died from respiratory distress during severe COVID-19 were tested for inflammatory infiltrates (Figure 2). Panel A shows extensive infiltration of broncho-pulmonary parenchyma by inflammatory cells with extensive damage of alveolar spaces, exfoliation of bronchial epithelium and thrombosis of blood vessels. Immunohistochemistry (Panel B) showed a prominent expression of granulocyte markers CD11b+ and CD66b+, with few CD68+ macrophages and virtually no T cells (data not shown). Double immunofluorescence (Panel C) confirmed that most inflammatory cells co-expressed CD11b+CD66b+, and immunohistochemistry confirmed the presence of large numbers of Arg1^+^ cells (Panel D). Double immunofluorescence (Panel E) demonstrated a high expression of granular intracytoplasmic Arg1 in CD66b+ confirming them to be Arg^+^G-MDSC. Furthermore, staining with anti-NOX-1 and NOX-2 antibodies revealed high expression of these enzymes in clusters of inflammatory and exfoliated epithelial cells (Panel F). NOX-1 and NOX-2 enzymes catalyze the synthesis of reactive oxygen species (ROS). These results suggest that the massive infiltration of the lungs by Arg^+^G-MDSC simultaneously expressing NOX1/2, may inhibit T cells and caused endothelial cell dysfunction (through depletion of arginine, which is required for nitric oxide production and blood vessel tone), and directly damage alveolar epithelium through the release of ROS. This may in part explain the acute respiratory distress syndrome and coagulopathy frequently seen in severe COVID-19 patients. The high numbers of Arg^+^G-MDSC in circulation and lungs of COVID-19 patients, contrasts with the findings in cancer where G-MDSC concentrate around tumors with increases in peripheral blood found mostly in patients with advanced disease^17-19^.

**Figure 2.**
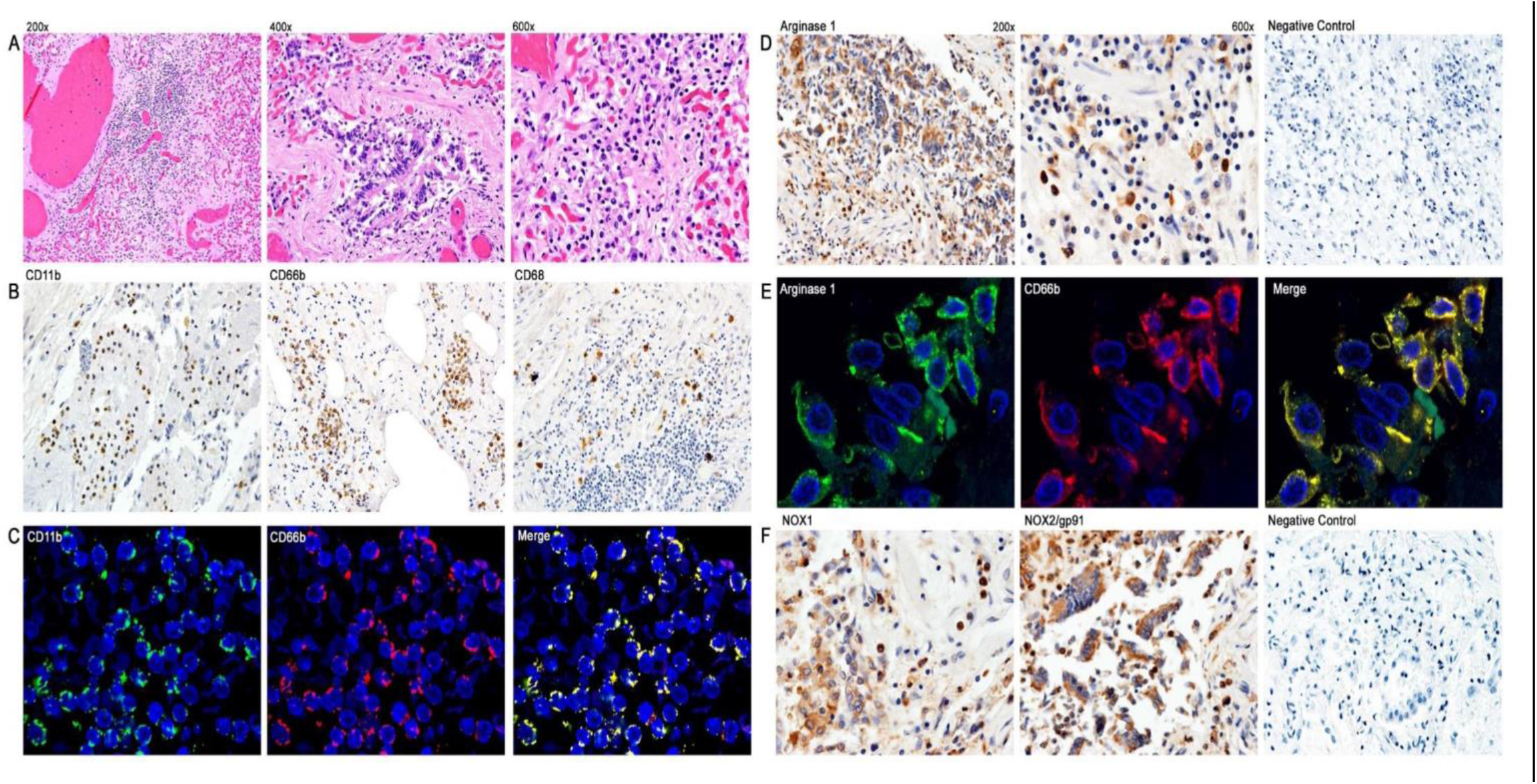
Immunohistochemistry and Double Immunofluoresence of Inflammatory Infiltrates in Lung Autopsy Samples. **A)** Hematoxylin & Eosin staining. **B)** Immunohistochemistry with anti-CD11b, CD66b and CD68. C) Double immunofluorescence with anti-CD11b and anti-CD66b. **D)** Immunohistochemistry for Arginase 1. **E)** Double immunofluorescence with anti-Arginase 1 and anti-CD66b; **F)** Immunohistochemistry with anti-NOX1 and anti-NOX2.

### Metabolic and functional consequences of increased Arg^+^G-MDSC

G-MDSC can deplete arginine and impair T cell and endothelial cell function^16 20,21^. Figure 3A shows representative Western blot data from 7 severe and 7 convalescent COVID-19 patients and 3 healthy controls. All severe COVID-19 patients had high Arg1 protein expression, compared to only 2/7 convalescent patients tested and none of the healthy controls. Quantification of these Arg1 expression levels reveals a roughly 5-fold increase in Arg1 in severe COVID-19 patients compared to controls and convalescent individuals. SYBR Green qRT-PCR using an MDSC primer panel confirmed a 7.5 fold higher expression of Arg1 in the PBMC of severe COVID-19 patients compared to convalescent and healthy controls (Figure 3B). These results also showed increased expression of genes associated with MDSC including MMP9, S100A9, CEBP and genes associated with granulocyte functions such as myeloperoxidase (MPO) and neutrophil degranulation proteins PRTN3. These findings were further confirmed by RNAseq from purified G-MDSC (shown in Figure 4)

**Figure 3.**
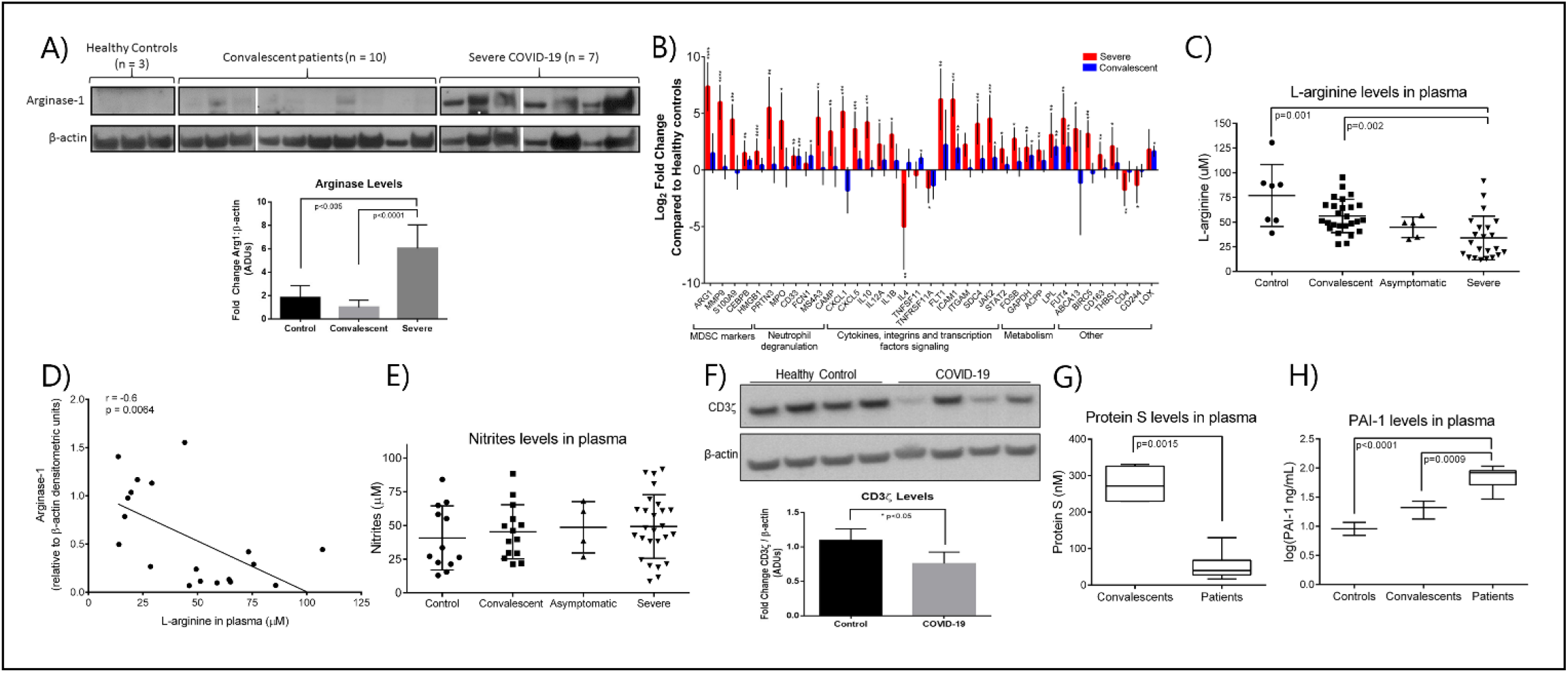
Effects of Increased Arg^+^G-MDSC. **A)** Arginase-1 protein expression in PBMC from severe, convalescent COVID-19 patients and healthy controls with associated quantification of fold-changes. **B)** Gene expression comparison by SYBR green qRT-PCR of PBMC from severe (red bars) and convalescent (blue bars) and healthy controls (baseline). X axis shows Log2 fold change in gene expression. Asterisks denote *p ≤ 0.05, **p ≤ 0.01, ***p ≤ 0.001, and ****p ≤ 0.0001, using Student’s t-test from the average ΔCt values. **C)** L-arginine concentration in plasma of patients and healthy controls **D)** Correlation of Arg-1 protein expression in PBMC and plasma arginine. **E)** Plasma nitrate levels in plasma. **F)** CD3ζ chain expression in purified T cells from 4 representative healthy controls and 4 COVID-19 patients with average densitometry shown below **G)** Plasma protein S and Plasminogen activator inhibitor-1 (PAI-1) in plasma.

**Figure 4.**
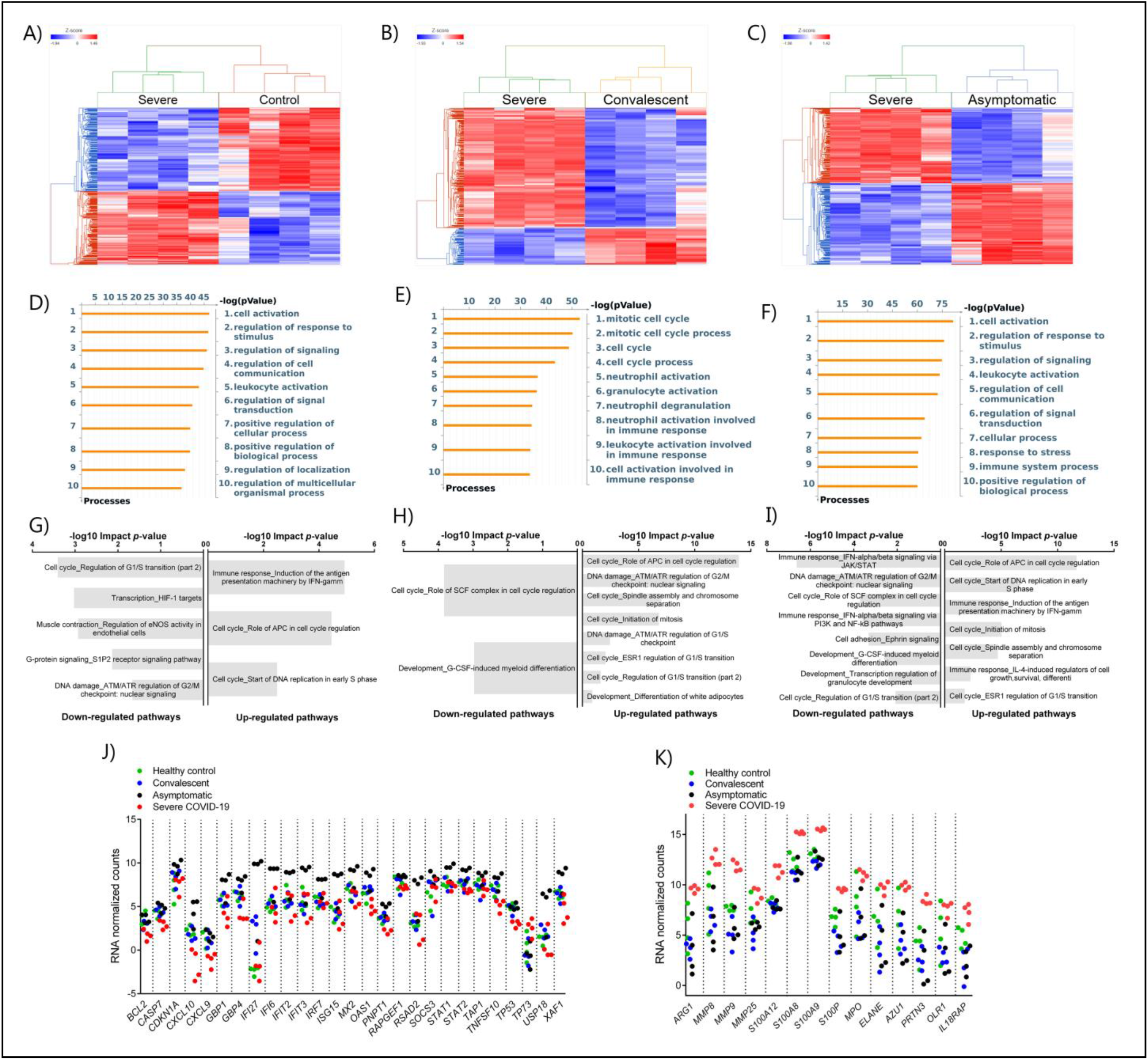
Differential Gene Expression in G-MDSC from Patients and Healthy Controls. Comparison of the transcriptome of purified G-MDSC using RNAseq from severe (n=4), asymptomatic (n=4), and convalescent (n=4) COVID-19 patients and healthy controls (n=4). **A-C)** Dendrograms and Heat-Maps for Unsupervised Hierarchical Clustering comparing transcriptome expression. **D-F)** Differences in Gene Ontology Processes identified by *MetaCore* in G-MDSC from patients and healthy controls. **G-I)** Analysis using Key Pathway Advisor software identified the Top 25 differentially expressed pathways. **J**,**K)** Dot plots comparing differentially expressed genes of “Immune response IFNalpha/beta signaling via JAK/STAT” (**J**) and Granulocyte Functions and Degranulation (**K**) in G-MDSC from patients and healthy controls.

The metabolic consequence of high numbers of Arg^+^G-MDSC was a significant decrease in plasma arginine in severe COVID-19 patients (mean=45µM; range 18-120µM), compared to healthy controls (mean=75µM; range 50-130µM) and convalescent patients (mean=55µM; range 26-92) (Figure 3C). As expected, there was an inverse correlation between Arg1 expression (by W. blot) and arginine plasma levels (Figure 3D), but nitrite levels were unchanged (Figure 3E). These observations are significant in that T cells cultured in arginine <50µM lose the T cell receptor zeta chain (CD3ς) expression, impairing proliferation and IFNγ production ^13,20^. This effect was also observed in purified T cells from severe COVID-19 patients tested (Figure 3F). Arginine depletion also causes endothelial cell dysfunction through interfering with nitric oxide production ^16^. This was confirmed here by decreased levels of Protein S and increased plasminogen activator inhibitor-1 (PAI-1)(Figure G), which can increase the risk for intravascular coagulation.

#### G-MDSC from COVID-19 Patients have Stage-Specific RNA Expression Profiles

To better understand the differences in inflammatory mechanisms in patients with different stages and severity of COVID-19, we compared the transcriptome of purified G-MDSC using RNAseq. Unsupervised hierarchical cluster analysis of the differentially expressed genes (DEGs) in G-MDSC from severe COVID-19 patients vs healthy controls (Figure 4A), convalescent (Figure 4B) and asymptomatic patients (Figure 4C) demonstrated clear differences between these groups. A Venn diagram (Supplemental Figure 1) illustrating shared and unique genes revealed the most significant differences in RNA transcripts (FDR <0.05; fold change ≥ 2) was between severe and asymptomatic patients (3675 transcripts), followed by severe and healthy controls (2452 RNA transcripts) and severe and convalescent (863 genes). An initial analysis using *MetaCore* software identified gene ontology processes that were significantly different among the groups. Specifically, G-MDSC from severe vs convalescent patients showed major differences in the expression of genes associated granulocyte functions and degranulation (Figure 4E). In contrast, differences between severe COVID-19 patients and healthy controls or asymptomatic patients were primarily associated with normal cell activation, signaling and regulation (Figure 4 D, E). Additional analysis using *Key Pathway Advisor* software identified the top 25 Pathways with predictive positive or negative functional consequences. Comparison of severe vs asymptomatic patients showed that G-MDSC from severe patients had significantly decreased expression of genes from pathways associated with type I IFN responses (Immune response_IFN-alpha/beta signaling via JAK/STAT) and its down-stream signaling mechanisms (Immune response_IFN alpha/beta signaling via PI3K and NF-ĸB pathways) (Figure 4I). To further illustrate these findings we plotted the expression of representative genes associated with granulocyte functions and genes associated with type I IFN pathways. These data confirmed increased expression of genes associated with anti-viral responses such as IFI27, IFIT2 and IFIT3 ^22-25^ in G-MDSC from asymptomatic patients(Figure 4J). In contrast, G-MDSC from severe COVID-19 patients had increased expression of genes associated with granulocyte functions/degranulation and chronic inflammation including Arg1, MMP8, MMP9, S100A8 and A9, MPO, IL18RA, elastase, PRTN3 (azurophilic granule protein 7) (Figure 4K). These results demonstrate contrasting inflammatory and immune responses between asymptomatic and severe COVID-19 patients. The increased expression of type I IFN associated pathways and down-stream signaling mechanisms in G-MDSC from asymptomatic patients suggested the development of a protective anti-viral immune response. It is unclear whether G-MDSC can themselves have anti-viral activity or serve as effective antigen presenting cells. In contrast patients with severe COVID-19 develop a granulocyte inflammatory response that exacerbates the disease. What genomic characteristics regulate the degree and type of inflammatory response is still unclear, but these findings may help identify individuals that are more likely to develop severe disease. The complete lists of differentially expressed genes are included in Supplemental Table 2.

## DISCUSSION

These data demonstrates the importance of Arg^+^G-MDSC in the pathophysiology of severe COVID-19. As previously reported arginine depletion impairs T cell and endothelial cell function. Here we further demonstrate the infiltration of lungs by Arg^+^G-MDSC expressing NOX1/NOX2, possibly release ROS causing direct damage of the pulmonary epithelium and increase inflammation, resulting in acute respiratory distress. Why G-MDSC show a preference for bronchoalveolar tissues and not other organs (data not shown) is unclear. It is possible that release of damage associated molecular patterns (DAMPS) from infected respiratory epithelium activate G-MDSC. Similar observations have been made in patients with COPD where DAMPS increase inflammation and fibrosis^26,27^. Alternatively lipopolysaccharides (LPS) from bacterial pneumonia, which frequently accompanies severe COVID-19, could exacerbate the response by G-MDSC. Severe alveolar damage caused by inflammatory cytokines and ROS has been well documented as a mechanism for pathogenesis of acute lung injury and adult respiratory distress syndrome^28^. Patients with Severe COVID-19 studied here also had significantly increased levels of inflammatory cytokines in plasma (Supplemental Figure 2).

Arginine depletion can also be a mechanism for endothelial cell dysfunction and increased intravascular coagulation. Inhibition of arginase-1 restores endothelial function and production of nitric oxide^14,15,29,30^. Our data shows that severe COVID-19 patients had decreased plasma levels of Protein S, suggesting it is rapidly being consumed, and increased PAI-1, which is a direct manifestation of endothelial cell dysfunction. Arginase can also decrease NO production by endothelial cells because of its faster kinetics and higher avidity for arginine compared to nitric oxide synthase 2 (NOS2). Thus, arginine deficiency can cause vasoconstriction, increase platelet adherence, and further promote hypercoagulation^30^.

These observations also suggest novel therapeutic approaches including the use of arginase inhibitors or the replenishment of arginine. Arg1 inhibitors are currently in early phase clinical trials in cancer (Calithera/Incyte; ^31,32^), while the replenishment of arginine or arginine precursors has been tested in Sickle cell disease resulting in a significant reduction in vaso-occlusive complications^33,34^ Arginine replenishment has also been tested in surgery where it successfully blunted the surge of G-MDSC, prevented T cell dysfunction and decreased infectious complications ^35-37^. Therefore arginase 1 inhibition and/or arginine replenishment should be considered as an adjuvant to the prevention/treatment of COVID-19.

### Methods

Detailed methods can be found in the supplemental data file

## Data Availability

Relevant data is all included in the body and supplemental information.

## Author Constributions

MD, MSP, RT, DW, PP: Processed samples, conducted flow cytometry, W blots, HPLC and qPCR

LDV, LD, RVH: Immunohistology

JZ, JG: RNA seq - transcriptome and analysis

JBO, JGD, LB, RN, WMR: Identified and consented patients, collected samples and clinical data

RM: Endothelial dysfunction

AC, JC, BL, RJ: Statistical analysis

CAR, CRM, HK: Provided critical input on data interpretation, new assays and manuscript development

ACO: Overall planning and execution, data interpretation, manuscript preparation

## Acknowledgments

This project was supported by funding from LSU Health and Ochsner Medical Center. Core facilities, personnel and services partially supported by *P20-GM103501, P20-CA233374 and P20-GM121288*. We would like to acknowledge the dedication of doctors, nurses and researchers at LSU Health and Ochsner Medical Center who provide care and research for COVID-19 patients, and to patients who participated in this study.

## References

1. Pedersen, S.F. & Ho, Y.C. SARS-CoV-2: a storm is raging. J Clin Invest 130, 2202–2205 (2020).

2. Wang, F., et al. The laboratory tests and host immunity of COVID-19 patients with different severity of illness. JCI Insight 5(2020).

3. Didangelos, A. COVID-19 Hyperinflammation: What about Neutrophils? mSphere 5(2020).

4. Chan, A.S. & Rout, A. Use of Neutrophil-to-Lymphocyte and Platelet-to-Lymphocyte Ratios in COVID-19. J Clin Med Res 12, 448–453 (2020).

5. Liu, Y., et al. Neutrophil-to-lymphocyte ratio as an independent risk factor for mortality in hospitalized patients with COVID-19. J Infect 81, e6–e12 (2020).

6. Agrati, C., et al. Expansion of myeloid-derived suppressor cells in patients with severe coronavirus disease (COVID-19). Cell Death Differ 27, 3196–3207 (2020).

7. Reizine, F., et al. SARS-CoV-2-Induced ARDS Associates with MDSC Expansion, Lymphocyte Dysfunction, and Arginine Shortage. J Clin Immunol (2021).

8. Falck-Jones, S., et al. Functional monocytic myeloid-derived suppressor cells increase in blood but not airways and predict COVID-19 severity. J Clin Invest (2021).

9. Gabrilovich, D.I. Myeloid-Derived Suppressor Cells. Cancer Immunol Res 5, 3–8 (2017).

10. Marvel, D. & Gabrilovich, D.I. Myeloid-derived suppressor cells in the tumor microenvironment: expect the unexpected. J Clin Invest 125, 3356–3364 (2015).

11. Bryk, J.A., et al. Nature of myeloid cells expressing arginase 1 in peripheral blood after trauma. J Trauma 68, 843–852 (2010).

12. Rodriguez, P.C., et al. L-arginine deprivation regulates cyclin D3 mRNA stability in human T cells by controlling HuR expression. J Immunol 185, 5198–5204 (2010).

13. Rodriguez, P.C. & Ochoa, A.C. Arginine regulation by myeloid derived suppressor cells and tolerance in cancer: mechanisms and therapeutic perspectives. Immunol Rev 222, 180–191 (2008).

14. Mahdi, A., Kövamees, O. & Pernow, J. Improvement in endothelial function in cardiovascular disease - Is arginase the target? International Journal of Cardiology 301, 207–214 (2020).

15. Masi, S., et al. Aging Modulates the Influence of Arginase on Endothelial Dysfunction in Obesity. Arteriosclerosis, Thrombosis, and Vascular Biology 38, 2474–2483 (2018).

16. Lucas, R., et al. Arginase 1: an unexpected mediator of pulmonary capillary barrier dysfunction in models of acute lung injury. Front Immunol 4, 228 (2013).

17. Kumar, V., Patel, S., Tcyganov, E. & Gabrilovich, D.I. The Nature of Myeloid-Derived Suppressor Cells in the Tumor Microenvironment. Trends Immunol 37, 208–220 (2016).

18. Lu, T., et al. Tumor-infiltrating myeloid cells induce tumor cell resistance to cytotoxic T cells in mice. J Clin Invest 121, 4015–4029 (2011).

19. Wesolowski, R., et al. Circulating myeloid-derived suppressor cells increase in patients undergoing neo-adjuvant chemotherapy for breast cancer. Cancer Immunology, Immunotherapy 66, 1437–1447 (2017).

20. Rodriguez, P.C., et al. Arginase I-producing myeloid-derived suppressor cells in renal cell carcinoma are a subpopulation of activated granulocytes. Cancer Res 69, 1553–1560 (2009).

21. Rodriguez, P.C., Ochoa, A.C. & Al-Khami, A.A. Arginine Metabolism in Myeloid Cells Shapes Innate and Adaptive Immunity. Front Immunol 8, 93 (2017).

22. Ramilo, O., et al. Gene expression patterns in blood leukocytes discriminate patients with acute infections. Blood 109, 2066–2077 (2007).

23. Tang, B.M., et al. A novel immune biomarker IFI27 discriminates between influenza and bacteria in patients with suspected respiratory infection. Eur Respir J 49(2017).

24. Tran, V., et al. Influenza virus repurposes the antiviral protein IFIT2 to promote translation of viral mRNAs. Nat Microbiol 5, 1490–1503 (2020).

25. Tretina, K., Park, E.S., Maminska, A. & MacMicking, J.D. Interferon-induced guanylate-binding proteins: Guardians of host defense in health and disease. J Exp Med 216, 482–500 (2019).

26. Pouwels, S.D., et al. Susceptibility for cigarette smoke-induced DAMP release and DAMP-induced inflammation in COPD. Am J Physiol Lung Cell Mol Physiol 311, L881–L892 (2016).

27. Pouwels, S.D., et al. Increased neutrophil expression of pattern recognition receptors during COPD exacerbations. Respirology 22, 401–404 (2017).

28. Kellner, M., et al. ROS Signaling in the Pathogenesis of Acute Lung Injury (ALI) and Acute Respiratory Distress Syndrome (ARDS). in Pulmonary Vasculature Redox Signaling in Health and Disease (ed. Wang, Y.-X.) 105–137 (Springer International Publishing, Cham, 2017).

29. Caldwell, R.W., Rodriguez, P.C., Toque, H.A., Narayanan, S.P. & Caldwell, R.B. Arginase: A Multifaceted Enzyme Important in Health and Disease. Physiol Rev 98, 641–665 (2018).

30. Kim, J.H., et al. Arginase inhibition restores NOS coupling and reverses endothelial dysfunction and vascular stiffness in old rats. J Appl Physiol (1985) 107, 1249–1257 (2009).

31. Borek, B., Gajda, T., Golebiowski, A. & Blaszczyk, R. Boronic acid-based arginase inhibitors in cancer immunotherapy. Bioorg Med Chem 28, 115658 (2020).

32. Grzywa, T.M., et al. Myeloid Cell-Derived Arginase in Cancer Immune Response. Front Immunol 11, 938 (2020).

33. Morris, C.R., et al. A randomized, placebo-controlled trial of arginine therapy for the treatment of children with sickle cell disease hospitalized with vaso-occlusive pain episodes. Haematologica 98, 1375–1382 (2013).

34. Onalo, R., et al. Randomized control trial of oral arginine therapy for children with sickle cell anemia hospitalized for pain in Nigeria. Am J Hematol 96, 89–97 (2021).

35. Braga, M., Gianotti, L., Vignali, A. & Carlo, V.D. Preoperative oral arginine and n-3 fatty acid supplementation improves the immunometabolic host response and outcome after colorectal resection for cancer. Surgery 132, 805–814 (2002).

36. Hamilton-Reeves, J.M., et al. Perioperative Immunonutrition Modulates Inflammatory Response after Radical Cystectomy: Results of a Pilot Randomized Controlled Clinical Trial. J Urol 200, 292–301 (2018).

37. Popovic, P.J., Zeh, H.J., 3rd & Ochoa, J.B. Arginine and immunity. J Nutr 137, 1681S–1686S (2007).

