## Supplemental Data for "Transcriptome and Functions of Granulocytic Myeloid-Derived Suppressor Cells Determine their Association with Disease Severity of COVID-19"

### I. METHODS

Patient Selection: The study was conducted under LSU IRB protocol #20-053 and Ochsner Medical Center IRB protocol 21015-101C. All participants were consented prior to inclusion in the study. The study included 24 severe, 5 asymptomatic, 26 convalescent COVID-19 patients and 15 healthy controls. All patients tested positive for SARS-CoV-2 by PCR. Severe patients were hospitalized while asymptomatic and convalescent patients remained outpatient. Healthy controls were negative either by PCR or for antibodies to SARS-Cov-2. Classification of the severity of COVID-19 was done using CDC criteria. All severe patients in this study were hospitalized in the intensive care unit for treatment because of hypoxia and respiratory distress or complications from pre-existing comorbidities. Autopsy samples from ten (10) patients who died from COVID-19 complications were collected at the LSU Health Science Center Pathology Department.

Sample processing: EDTA and sodium citrate anticoagulated peripheral blood samples were centrifuged for separation of plasma and cellular components. Plasma was frozen at -70$^{o}$C and buffy coat was overlaid on ficoll-hypaque for separation of peripheral blood mononuclear cells (PBMC).

Flow cytometry Briefly, whole blood or PBMC were stained for CD45 to identify immune cells, and antibodies against T cells (CD3, CD4 and CD8), NK cells (CD16), monocytes (CD68) and MDSC subpopulations. The latter were divided into G-MDSC ($CD11b^{+}CD66b^{+}CD14^{-}$), and monocytic or M-MDSC ($CD11b^{+}$ $CD14^{+}HLA\text{-}DR^{low/neg}$).

Histology and Immunohistochemistry: Immunohistochemistry was performed using the avidin-biotin-peroxidase methodology, according to the manufacturer's instructions (Vectastain ABC Elite Kit, Vector Laboratories, Burlingame, CA). Our modified protocol includes deparaffination in xylenes, rehydration through descending grades of ethanol up to water, non-enzymatic antigen retrieval with 0.01 M sodium citrate buffer pH 6.0 heated to 95°C for 25 minutes in a vacuum oven, endogenous peroxidase quenching with 3% H2O2 in methanol, blocking for 2 hours with normal horse serum (for mouse monoclonal antibodies) or normal goat serum (for rabbit polyclonal or recombinant rabbit monoclonal antibodies) and incubation with primary antibodies overnight at room temperature in a humidified chamber. After rinsing in PBS, sections were incubated with biotinylated secondary antibodies for 1 hour at room temperature, followed by incubation with avidin-biotin-peroxidase complexes for 1 hour. Finally, the peroxidase was developed with diaminobenzidine (Boehringer, Mannheim, Germany) for 3 minutes, and the sections were counterstained with Hematoxylin and mounted with Permount (Fisher Scientific, Pittsburgh PA). Antibodies used in this study for the characterization of immune cells included: a CD3 mouse monoclonal (Clone F7.2.38, 1:100 dilution, DAKO-Agilent Technologies, Santa Clara, CA), a CD20 mouse monoclonal Clone L26, 1:100 dilution, DAKO), a CD11b rabbit monoclonal, raised against the C-terminal (Clone EP1345Y, 1:100 dilution, Abcam, Cambridge, MA), a CD66b mouse monoclonal (Clone 80H3, 1:100 dilution, LifeSpan Biosciences, Seattle, WA), and a CD68 mouse monoclonal (Clone PG-M1, 1:100 dilution, DAKO). Other antibodies included a rabbit polyclonal anti-Arginase-1 (H-52, 1:500 dilution, Santa Cruz Biotechnology, Dallas, TX), a rabbit polyclonal anti-Nox1 (ab78016, 1:500 dilution, Abcam), and a mouse monoclonal anti-NOX2/gp91phox (Clone 54.1, 1:200 dilution, Abcam). Bright field photomicrographs were taken with an Olympus DP72 Digital Camera using an Olympus BX70 microscope (Olympus, Center Valley, PA).

Double Labeling Immunofluorescence. Deparaffination and rehydration of tissues were performed as described above. Antigen retrieval was also performed with heat and citrate buffer; however, no endogenous peroxidase quenching is necessary. After overnight incubation with the first primary antibody (mouse or rabbit), sections were rinsed with PBS, and an AlexaFluor-568-tagged secondary antibody was incubated for 2 hours at room temperature in the dark. After thoroughly washing with PBS three times, a second primary antibody (raised in a different species than the first) was incubated overnight at room temperature in a dark humidified chamber. Finally, a second AlexaFluor-488-conjugated secondary antibody was incubated for 2 hours at room temperature in the dark, and finally, after rinsing in PBS, sections were coverslipped with an aqueous-based mounting media (Vectashield® Hard Set with DAPI; Vector Laboratories), and visualized in an Olympus FV1000 confocal microscope equipped with FluoView software. Confocal scanning of double labeled sides was done with the "Sequential" and Kalman imaging functions of the confocal, which prevents bleaching of the fluorescent signal through different channels and to eliminate background signal, ensuring the accuracy of the images.

Double Labeling Immunofluorescence. After overnight incubation with the first primary antibody  sections were rinsed and an AlexaFluor-568-tagged secondary antibody was incubated for 2 hours at room temperature in the dark. After thorough washing, a second primary antibody (raised in a different species than the first) was incubated overnight at room temperature in a dark humidified chamber. Finally, a second AlexaFluor-488-conjugated secondary antibody was incubated for 2 hours at room temperature in the dark. Sections were coverslipped with an aqueous-based mounting media (Vectashield® Hard Set with DAPI; Vector Laboratories), and visualized in an Olympus FV1000 confocal microscope equipped with FluoView software. Confocal scanning of double labeled sides was done with the "Sequential" and Kalman imaging functions of the confocal, which prevents bleaching of the fluorescent signal and eliminated background signals, ensuring the accuracy of the images.

Western blot for Arginase1 in G-MDSC and CD3ζ chain in T cells. Protein extract was obtained by lysing PBMC or purified CD3 cells using Mammalian Protein Extraction Reagent (Thermo Scientific) supplemented with 0.1% SDS and Halt Protease and Phosphatase Inhibitor. Twenty μg (20μg) of protein was loaded into an 8% Bis-tris gel (Life Technologies) for detection of Arg1, or a 4-12% Bis-tris gel for detection of CD3ζ.  After transfer to a PVDF membrane it was blocked with 5% milk for 1h and incubated overnight with appropriate primary antibodies.  Primary anti-human antibodies included arginase-1 (BD Biosciences), phospho CD3ζ (Abcam), CD3ζ (Abcam), and β-actin. Following incubation HRP was detected using ECL Western Blotting Substrate (Thermo Scientific).

Gene Expression:  Gene expression in PBMC from COVID-19 patients and healthy controls was tested by qRT-PCR using a pre-designed Human Myeloid Derived Suppressor Cell Primer Library from RealTimePrimers.com. Briefly PBMC from healthy controls (n=5), severe (n=6), and convalescent COVID-19 patients (n=6) were centrifuged and RNA was extracted using Qiagen RNeasy Mini extraction kit and used for cDNA synthesis (Thermo Verso cDNA synthesis kit). Amplification by SYBR Green based qRT-PCR Reactions were conducted in duplicate and Ct value technical replicates were averaged. ΔCt values of each gene were calculated by normalizing gene expression against the housekeeping control gene PPIA. $2^{-\Delta\Delta Ct}$ was calculated to determine the gene expression fold change compared to healthy controls, and expressed as the log2 fold change. Error in gene expression was included as the standard deviation of the ΔΔCt. Statistical significance was determined by calculating the p-value using a student's t-test from the average ΔCt values of either active or convalescent patients compared to control patients. A fold change of an increase of 2 or less than 0.5 was also used as a statistical threshold. Only genes that provided significance in both methods were reported.

Plasma Arginine was measured high-performance liquid chromatography (HPLC) electrochemical detector (E1 = 150, E2 = 650mV) using a ThermoScientific Dionex Ultimate 3000. Standards of L-arginine were run with each experiment and the levels of the amino acid in a test sample were determined relative to this standard curve. RPMI media (1.15mM L-arginine) was used as an internal control.

Cytokines, Nitrites, Protein S and PAI-1 inhibitor: Plasma was used to measure cytokines using a Multiplex assay (Milliplex MAP Human Cytokine/Chemokine Magnetic Bead Panel Millipore-Sigma). Nitrites formed by the spontaneous oxidation of NO were measured by Griess reaction (Molecular Probes). Endothelial cell dysfunction markers were measured by ELISA, namely changes in Protein S (Diapharma Inc. West Chester, OH) and PAI-1 Inhibitor (PAI-1- Abcam, Cambridge, MA).

RNA Sequencing: RNA sequencing and analysis was done at the Stanley S. Scott Cancer Center's Translational Genomics Core (TGC; LSUHSC, New Orleans, LA). Briefly, purified G-MDSC were isolated by labeling with anti-CD66b-PE (Clone G10F5; BD Biosciences) followed by positive selection with Human PE Positive Selection kit containing dextran-coated magnetic particles (Stemcell Technologies, Cambridge, MA). The CD66b-PE depleted fraction was used to isolate $CD3^+$ T cells by using the T cell negative isolation kit (Stemcell Technologies, Cambridge, MA). Total RNA was extracted from enriched cells by using Qiazol followed by purification with miRNeasy Mini Kit (both from Qiagen, Germany) with an additional DNase I treatment. RNA concentration was determined by fluorometry using the Qubit RNA HS Assay kit (Invitrogen, Thermo Fisher Scientific) and RNA integrity was assessed using the RNA 6000 Pico Kit on an Agilent Bio Analyzer 2100 (Agilent Technologies, Santa Clara, CA). Only samples that showed satisfactory amount and quality were used for RNA-Seq. Paired-end libraries (2 x 75) were prepared (600ng per sample) using the mRNA Stranded Library Preparation Kit from Illumina (Illumina Inc., San Diego, CA). The libraries were validated, normalized and sequenced on a NextSeq 500/550 High Output Kit v2.5 flow cell (Illumina) on the NextSeq500 sequencer (Illumina). FASTQ output files were downloaded from the Illumina BaseSpace and uploaded to Partek Flow. Contaminants (rDNA, tRNA, mtrDNA) were removed using Bowtie2 (version 2.2.5) and the unaligned reads were aligned to STAR (version 2.6.1d) using the hg38 version of the human genome as a reference. Aligned reads were quantified to the hg38-RefSeqTranscript release 93 and normalized by TMM followed by transformation by log2 (1+TMM+log2). Normalized counts were used to determine differential gene expression between patients with or without COVID-19 by using DEseq2 and with a false discovery rate for multiple testing (FDR) of 0.05 (FDR <0.05) and fold change 2 in Partek Flow. Genes with an FDR < 0.05 and fold change (FC) $\geq$ 2 were considered differentially expressed between the groups. Significantly differentially expressed genes were plotted as heat-maps with hierarchical clustering using the embedded algorithm in Partek Flow for hierarchical unsupervised comparison of the samples using Euclidian distance. The values of expression are visualized with colors ranging from red (high expression) over black (intermediate expression) to blue (low expression). We used MetaCore and Key Pathway Advisor software to predict pathways, networks, gene ontology processes and diseases associated with the differentially expressed genes. Pathways with a positive and negative direction in the Top 25 were selected for further analysis and pathways with no predicted functional consequences were not included. The datasets (FASTQ files, raw counts) are being granted an accession number at the Gene Expression Omnibus (GEO) and will be open to the public upon acceptance of the manuscript.

Statistical Analysis:
Comparisons between groups were done by one-way ANOVA on ranks and Dunn's Multiple Comparison Test. Two-tailed statistical significance of the Spearman's coefficient was calculated for correlating variables. Results were considered significant when $p \leq 0.05$. All analyses were done using GraphPad Prism 6 software (Graphpad, San Diego, CA).

For further comparisons of the significance and predictive values of the flow cytometry results were calculated using a t-test comparing log-values in the severe group against all other groups combined, without assuming the variances are equal. The log10-scaled observations in figure 1H are smoothed using a Gaussian kernel. Statistics and Figure1H were generated using R 4.0.2 and the following packages: tidyverse (1.3.0) and MESS (0.5.7).Confidence intervals for the sensitivity, specificity, PPV and NPV, were determined by a Bootstrap sampling with replacement. Using the CD66b:CD3 ratios as an example, there are 24 severe and 44 non-severe ratios. Each Bootstrap trial randomly selects 24 values from observed ratios or levels for severe cases. Every time a value is selected, it is returned to the list of possible values so that it could be selected again. Similarly, 44 values would be selected from the observed ratios or levels from non-severe samples. Each selected value is compared to the optimum threshold for this dataset and a 2x2 contingency table is constructed. From this table the sum of the sensitivity and specificity, as well as the sum of the PPV and NPV are stored. This trial was repeated a total of 10,000 times. Since the optimum observed sums from the original data are quite large, the distribution of sums across the 10,000 Bootstrap trials is not expected to be normally distributed. Therefore, the 10,000 values of the sensitivity, specificity, PPV and NPV are independently sorted from lowest to highest and the 251$^{st}$ and 9750$^{th}$ values represent the lower and upper bound of the 95% confidence interval for each parameter.

To assess the significance of the observed sensitivity, specificity, PPV and NPV, permutation testing was used. In each trial, the ratios or levels of all samples were scrambled and them used to construct a 2x2 contingency table. After 10,000 trials, the number of times the calculated sensitivity, specificity, PPV, NPV, as well as the sum of the sensitivity and specificity, and the sum of the PPV and NPV exceed the observed values were used to determine the 1-tailed probability of observing these values by chance.

**SUPPLEMENTAL TABLE 1.**

Table 1. Demographics of COVID-19 Patients

| Participants | Number | Age (mean) | Sex | | Race | | | Ventilator Support** | | Body Mass Index Mean |
|---|---|---|---|---|---|---|---|---|---|---|
| | | | M | F | Caucasian | AA | Other | Yes | No | |
| **Severe** | 24 | 67.8 | 14 | 10 | 16 | 6 | 2 | 11 | 13 | 32.5 |
| **Asymptomatic** | 5 | 43.4 | 3 | 2 | 4 | 0 | 1 | No | | 27.0 |
| **Convalescent** | 26 | 49.3 | 13 | 13 | 22 | 4 | 0 | No | | 31.0 |
| **Normal Controls** | 15 | 53.5 | 11 | 4 | 11 | 0 | 4 | No | | 25.0 |

**Supplemental Figure 1.** A Venn diagram (Supplemental Figure 4G) illustrating the shared and unique genes among the groups

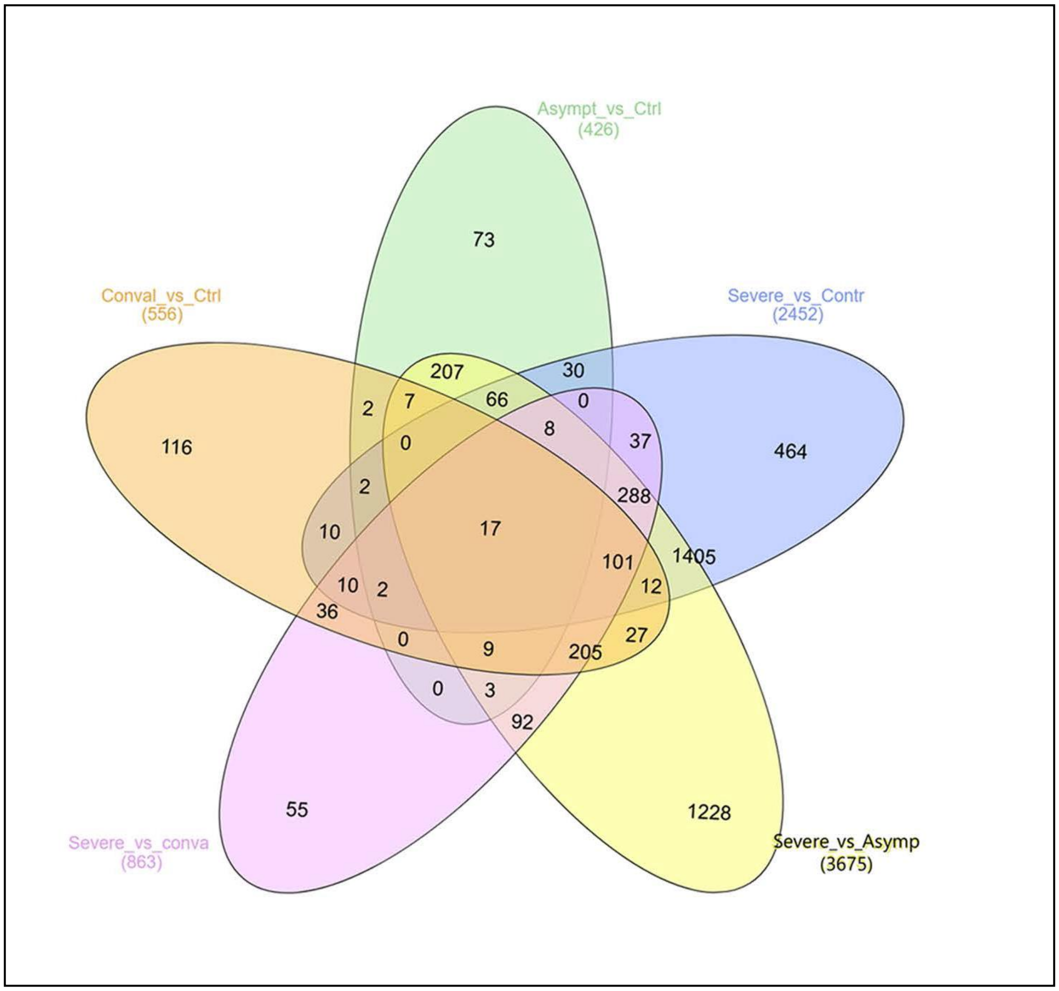

**SUPPLEMENTAL FIGURE 2.** Cytokines in plasma from COVID-19 patients were measured in Multiplex Assay and are expressed in pg/ml.

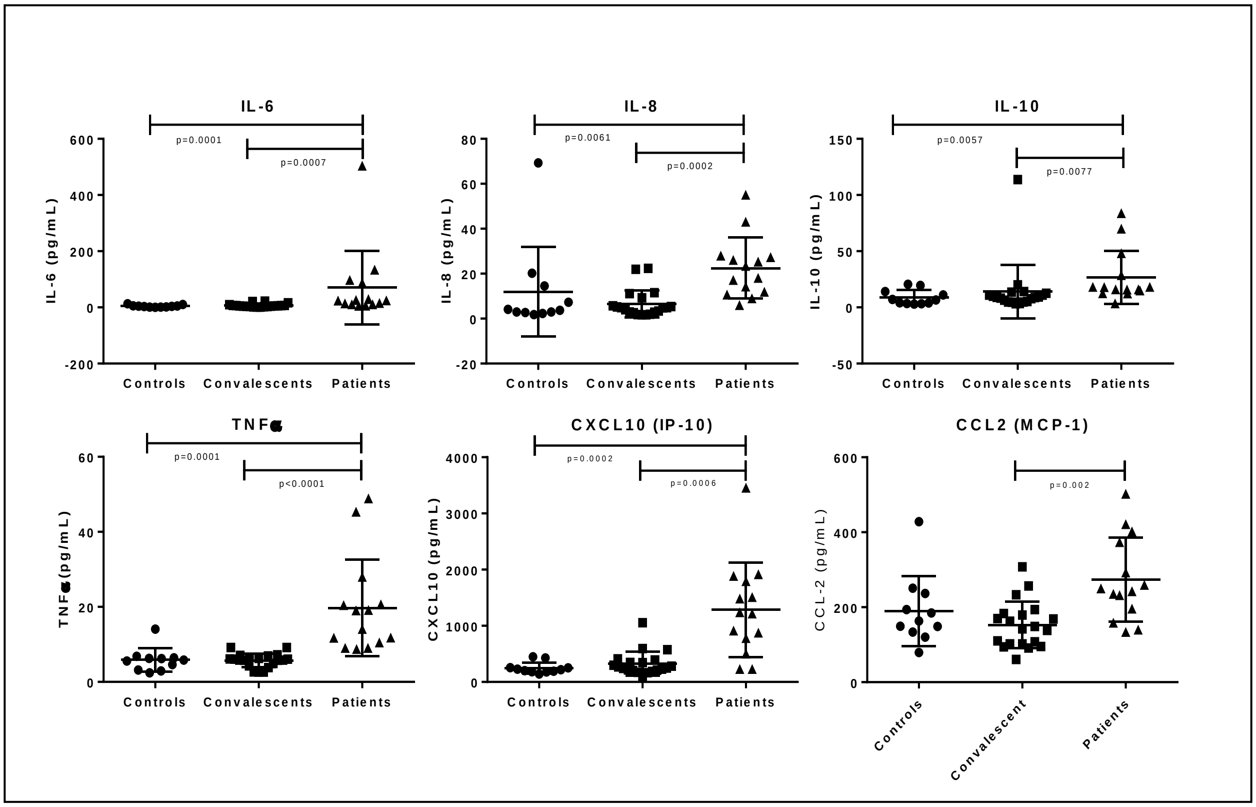
